# Neurophysiologic evidence for increased memory suppression among negative ruminators

**DOI:** 10.1101/2020.05.17.20104935

**Authors:** Aarti Nair, Joshua C. Eyer, Mark E. Faust

## Abstract

**Purpose:** Events (e.g., seeing a familiar face) may initiate retrieval of associated information (e.g., person’s name), but not all cue initiated memory retrieval is welcome (e.g., trauma). Memory suppression refers to the ability to halt unwanted retrieval, and any erosion of memory associations in response to repeatedly excluding a memory from consciousness. The current study sought to examine event related potential (ERP, averaged scalp electrical recordings) correlates of inhibitory cognitive control of memory retrieval and any linkage of such control to ruminative memory styles.

**Method:** Participants (N=23) first learned face-picture pairings. ERPs were then recorded as they viewed face cues while either bringing the associated picture to mind (think trial), or not allowing the associated picture to come to mind (no-think trial).

**Results:** Emotional valence of learned pictures (negative vs neutral) modulated a posterior (P1, 100-150 ms) ERP associated with attention to the face cue. Memory strategy (think vs no-think) modulated a frontal (P3, 350-450 ms) associated with alerting of the need to control retrieval. Both valence and strategy worked in combination to modulate a late posterior (LC, 450-550 ms) ERP associated with successful memory retrieval. Brooding, a negative form of rumination, was found to be positively correlated with the LC ERP.

**Conclusions:** The results suggest early separation of emotional and strategic control of retrieval, but later combined control over access to working memory. Moreover, the positive correlation of brooding and the LC suggest that individuals who are high in application of perseverative strategies to memory retrieval will show greater modulation of the retrieval-related LC ERP.

## 1 Introduction

Imagine that as you stand in line for a morning cup of coffee you realize the person in line in just front of you was someone you first met at a recent social gathering. Seeing this person cues retrieval of several memories from that gathering, but you are disappointed that memory of the person’s name is not recalled. You are further dismayed to find that memory of an embarrassing moment at the gathering you would rather not think about has been cued and retrieved. While much research effort has gone into studying successful cued recall, there is an increasing recognition that it is not uncommon for cues to initiate retrieval of memories we would rather not think about (Anderson & Levy, 2009; Levy & Anderson, 2002). Moreover, trauma and mood disorders often result in increased ruminations involving negative memories (Nolen-Hoeksema, 2000; Nolen-Hoeksema & Morrow, 1991).

### 1.1 Halting Retrieval and Memory Suppression (Think/No-think Paradigm)

To study this problem, Anderson and Green (2001) developed a procedure (think/nothink task) where participants first learned cue-target pairings to a criterion level, followed by a second phase where they were presented with a cue item and instructed to either allow the associated target item to come to mind (i.e., think trials) or to *not* allow the target item to come to mind (i.e., no-think trials). They reasoned that active cognitive control processes could (a) halt a cue-initiated retrieval (i.e., halting an unwanted retrieval), and (b) repeated successful disruption of retrieval could result in a weakening of the association between cue and target, making initiation of cued retrieval less likely for that cue-target pair in the future (i.e., memory suppression). Successful halting of cue-initiated retrieval, at least for some trials, can be indexed by comparing think and no-think cue-target pairs on a final cued recall test following the think/no-think phase of the procedure. A longer term memory suppression effect can be indexed by comparing final cued recall for no-think trials with baseline cue-target pairs that were learned to criterion, but were not included in the think/no-think phase. This distinction between halting and memory suppression effects is an important one, as halting effects are robust and have been widely replicated, but memory suppression effects for the think/no-think paradigm are not always present (Anderson & Levy, 2009; Bulevich, Roediger, Balota, & Butler, 2006). Memory suppression effects have been found to be sensitive to strategy differences during the think/nothink phase (van Schie, Geraerts & Anderson, 2013), and to memory style variables (e.g., rumination, Dieler, Herrmann & Fallgatter, 2014; Koster, De Lissnyder, Derakshan, & De Raedt, 2011).

However, while theoretical issues regarding long-term memory suppression effects are important for a better understanding of human memory (Anderson & Levy, 2007), short-term inhibitory cognitive control processes associated with halting unwanted memories are important also. Study of both short- and long-term memory suppression has relevance to clinical populations where ruminations on traumatic or negative memories play a role. Several studies have documented impaired memory suppression in depression (Hertel & Gerstle, 2003; Sacchet et al.,2017) and PTSD (Catarino, Kupper, Werner-Seidler, Dalgleish, Anderson, 2015; Hulbert & Anderson, 2018). Much of the early studies of the think/no-think task have used randomly paired words to form cue-target pairs that are learned during the initial phase of the task. To improve the external validity of the think/no-think task in applied settings, version of the task that use emotionally valenced target items, and that use visual objects and scenes to better model a common situation where a visual event cues retrieval of a visual memory.

This approach was adopted by Depue, Banich, and Curran (2006) who used the think/nothink paradigm to examine memory suppression for face-picture pairs where faces with a neutral expression were paired with photographs of emotionally valenced events selected from the International Affective Picture Series (IAPS; Lang, Bradley, and Cuthbert, 1995). Results demonstrated that memory suppression effects were greater for negatively valenced stimuli than neutrally valenced stimuli. Participants were able to recall the think-negative condition pairs better than the think-neutral condition pairs and were able to suppress the no-think-negative condition pairs better than the no-think-neutral condition pairs. Lambert et al. (2010) found a similar greater suppression effect for negatively valenced word pairs (e.g., cruel-socks) versus positively valenced word pairs (e.g., joy-socks).

### 1.2 Neural Basis of Memory Suppression

Studies using fMRI to image brain regions involved during performance of the think/nothink task suggest that prefrontal networks send control signals to regulate temporal lobe memory access and the activity of modality-specific cortical representations. Depue, Curran, and Banich (2007) scanned participants during the think/no-think phase using neutral faces paired with negatively valence pictures. They reported evidence for an early inferior frontal gyrus initiated regulation of hippocampus (memory retrieval) and amygdala (emotional content processing), and a slower medial frontal gyrus regulation of visual cortical areas supporting visual awareness of the retrieved target picture. They also suggested that the frontal poles may be involved in regulation of posterior cortical representations of emotional content of retrieved memory representations. Gagnepain, Hulbert, & Anderson (2017) extended this work. They used neutral face cues and both neutrally and negatively valenced target pictures and found evidence that dorsolateral frontal cortex regulated both hippocampus (and associated hippocampal cortex) and the amygdala using separate pathways. They verified frontal regulation of posterior visual cortex.

Previous electroencephalography (EEG) research on memory retrieval has identified that the correlate of a conscious successful recollection event is a positive shift of event-related potential (ERP) in parietal regions, typically left-lateralized and maximal approximately 400 to 800 ms after stimulus presentation (Rugg, 1995). This effect has also been found to be absent in patients with impaired recollection due to hippocampal lesions (Duzel, Vargha-Khadem, Heinze, & Mishkin, 2001), and it has been suggested to originate from recollection-related activity in hippocampal-parietal cortical networks (Curran, Tepe, and Piatt, 2006).

Bergstrom et al. (2007) provided the first ERP study of the think/no-think task (unrelated neutrally valenced word pairs). ERPs were recorded during the think/no-think phase (following initial learning of pairs to criterion). Both think and no-think ERP trials were segregated into recalled and unrecalled subcategories. This allowed them to generate ERP curves for the critical recalled-think trials to the unrecalled-think and both the recalled and unrecalled-no-think trials, thereby strengthening the interpretation of larger positive parietal component in a 500-800 ms time window (Late Component, LC) for the recalled-think trials (than for all the other trial types) as reflecting increased cortical processing of retrieved visual representations. Larger LC for think than for no-think trials has been repeatedly replicated (e.g., Bergstrom, Fockert, & Richardson-Klavehn, 2009, Hanslmayr, Leipold, Pastotter, & Bauml, 2009). Other studies identified earlier components leading up to the retrieval event that have been argued to be associated with cognitive control over memory retrieval. Several ERP studies have found evidence for specific early frontal components associated with memory suppression. The earliest is the P1, and early posterior positive component (100-150 ms) that is typically larger for think than for no-think trials and associated with attention to visual characteristics of the visual cue (Bergstrom et al., 2007; Chen et al., 2012). There is also typically a frontal N2 effect (a negative peak around 200 ms) that is more negative for no-think than for think trials, and has been associated with strategic cognitive control (Bergstrom et al., 2009, Chen et al., 2012). In a later time window, Hanslmayr et al. (2009) found a positive frontal P3-like effect (peaking ~300ms) with greater positivity for think than for no-think trials. The P3 positivity declined for no-think trials, but not for think trials across repetitions of cue-target pairs during the think/no-think phase. They argued that the frontal P3 was acting as an alerting signal for later cognitive control processes. Depue et al. (2013) additionally found increased theta power in frontal and parietal regions for items successfully suppressed after no-think trials compared to those that participants still remembered during the final recall phase. Our study was designed to assess the posterior P1, frontal N2, frontal P3, and posterior LC effects that have been associated with earlier cognitive control, and later retrieval success ERP signals.

### 1.3 Rumination and Memory Suppression

Rumination involves repetitive focusing on causes, situational factors, and consequences of negative life experiences (Nolen-Hoeksema & Morrow, 1991). An increased tendency towards rumination has been argued to be an important factor in depression and anxiety disorder (Notlen-Hoeksema, 2000), and evidence from large longitudinal studies of both adolecents and adults supports rumination as a transdiagnostic factor predicting the comorbidity of these mood disorders (McLaughlin & Nolen-Hoeksema, 2011). The view that rumination involves perseverative focus on certain memories, combined with the proposal that a breakdown in the ability to disengage attention from distracting or negative thoughts or information (Koster, Lissnyder, Derakshan, & De Raedt, 2011), suggests a relationship between memory suppression and rumination. Studies using the think/no-think pararadigm have found a negative correlation between rumination and percent recall on no-think trials (Hertel & Gerstle, 2003).

Because repeated repetition of think trials during performance of the think/no-think task may result in facilitation of final cued recall for think trials. By contrast, to the extent that halting retrieval of unwanted targets on no-think trials results in memory suppression, reduced final cued recall for think trials is expected. To be able to experimentally isolate the facilitative and suppression effects during the think/no-think task, it is customary to include baseline cue-target pairs initially learned to criterion but not included during the think/no-think phase of the te task. Rumination, has been found to be negatively correlated with memory suppression (indexed as baseline minus no-think recall, Fawcett et al., 2015). By contrast, a second study found that only a brooding (negative ruminations) was negatively correlated with memory suppression (baseline minus no-think). Several studies have documented impaired memory suppression in depression (Hertel & Gerstle, 2003; Sacchet et al.,2017) and PTSD (Catarino, Kupper, Werner-Seidler, Dalgleish, Anderson, 2015; Hulbert & Anderson, 2018).

### 1.4 Study Goals

Building on these reported findings, the current study sought to extend the literature by further documenting the ERP correlates of memory suppression during the think/no-think task specifically for valenced pictorial stimuli. To date, there are only 2 reports of think/no-think ERPs using both negative and neutral valenced picture targets (Chen, et al., 2012; Zhang et al., 2016). We wanted to verify the posterior P1 and LC, and the frontal N2, ERP components reported by these studies using emotionally valenced visual scenes. Moreover, we wanted to see if a frontal P3, an attentional anticipatory cognitive control signal reported by Hanslmayr et al. (2009) for neutral word stimuli would be found for emotionaly valences visual scenes. We are aware of no studies extant in the literature that have assessed the relationship of ERP markers of memory suppression during any version of the think/no-think task to rumination psychometric scores. Given that previous studies have found a negative correlation between rumination scores and cued recall for no-think trials and for baseline corrected no-think trials (Dieler et al., 2014; Fawcett tet al., 2015) we expected to find a negative relationship between rumination and the posterior LC ERP thought to index successful memory retrieval (Rugg, 1995).

## 2 Material and Methods

### 2.1 Design

The current study is a factorial memory experiment with measurement of EEG and cued recall responses measured for face-picture pairs. The design is a 2 emotional valence (neutral vs negative pictures) x 2 strategy (think vs no-think) completely repeated measures design.

Standard ANOVA models (extra factors are added to the ANOVA model for electrode locations where appropriate) are used to analyze memory effect in the recall and EEG results. This study followed a protocol approved by the University of North Carolina-Charlotte (UNCC) IRB, and written informed consent was obtained from all participants at time of testing.

### 2.2 Participants

Participants were college students recruited from the UNCC Department of Psychology Research Participation Pool. A total of 33 participants were recruited and received course credit for their participation. Of these, 10 were excluded due to equipment failure, excessive somnolence, noncompliance with task instructions, and/or high movement artifact, leaving a final sample of 23 participants. Two stimulus lists were created by counterbalancing pairing of each target with a randomly chosen male or female cue face across lists. Participants were pseudorandomly assigned to the 2 stimulus list conditions with the constraint of balances sample sizes (N=12, 11, for lists A & B). The mean age of the final sample was 25.3 (*SD* = 9.3). The sample was comprised of 18 females (78.3%) and 5 males (21.7%), and 73.9 % were Caucasian, 8.7 % were African-American, 4.3 % were Asian, and 13.1% were of other ethnicities. All participants were native speakers of English and right-handed, with normal or corrected-to-normal vision that allowed them to easily read text and clearly identify and describe pictures presented on the computer screen.

### 2.3 Material and Procedures

Participants were administered the Edinburgh Handedness Inventory (EHI; Oldfield, 1971) to control for handedness (only right-handers included). Participants were also administered the Ruminative Responses Scale (RRS; Nolen-Hoeksema & Morrow, 1991) which is a self-report measure of rumination that identifies two rumination sub-factors: reflection and brooding, the former represents adaptive rumination and the latter reflects maladaptive rumination (Treynor, Gonzalez, Nolen-Hoeksema, 2003).

The pictorial stimuli used for the Think/No-Think task (Figure 1) included 60 photographs of faces, half male and half female, validated to have neutral expressions by Depue et al. (2006). A separate 30 images with neutral emotional valence (e.g., pizza, freeway, baby) and 30 with negative emotional valence (e.g., funeral, scenes of war, electric chair) were also selected from the IAPS (Lang, Bradley, & Cuthbert, 2005). The faces and pictures were pseudorandomly paired to result in four groups of 15 stimuli blocked by sex (male/female) and valence (negative/neutral). Stimuli were presented using E-prime v1.1 software (Psychology Software tools, Pittsburgh, PA, USA).

**Figure 1.**
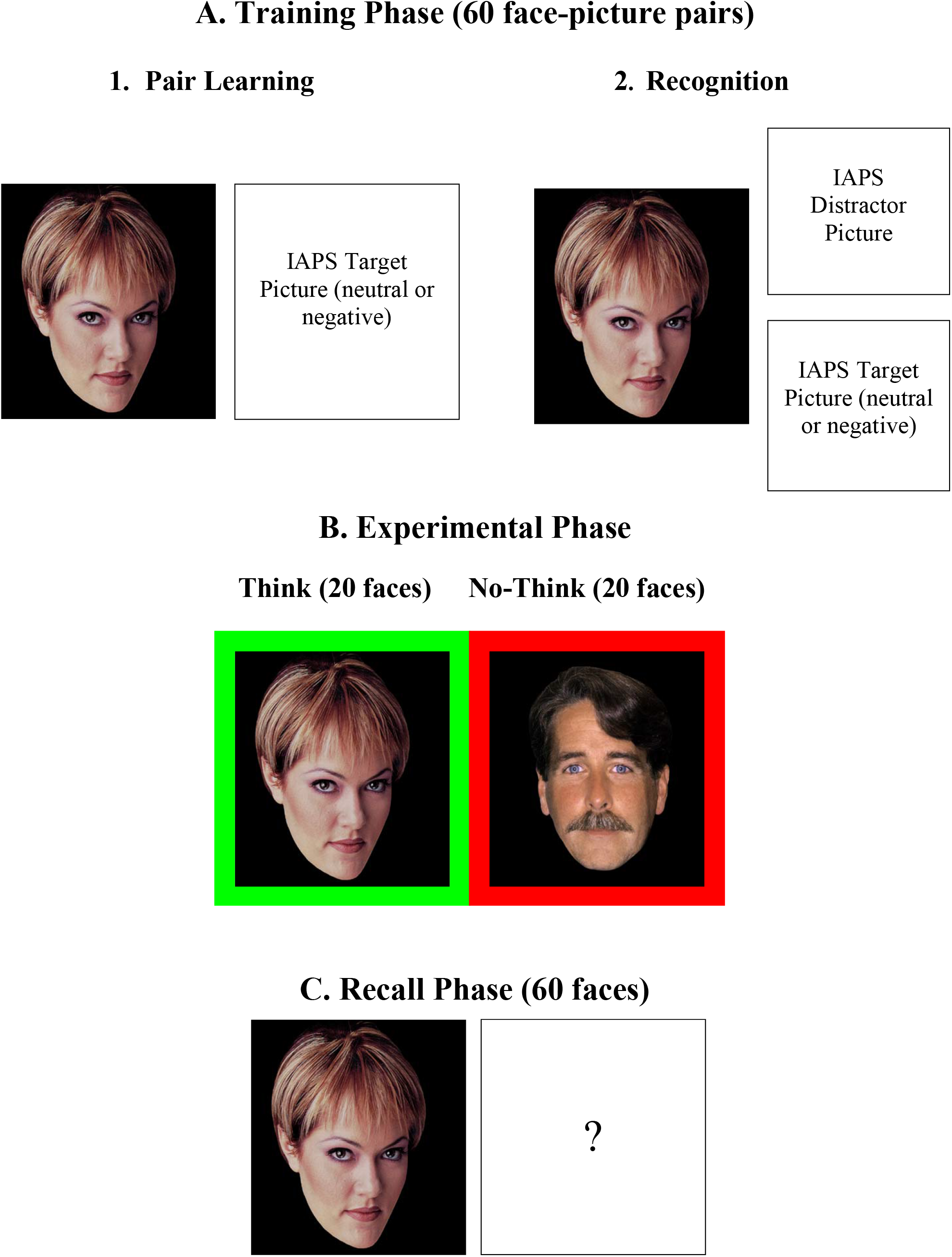
Think-No Think Paradigm using the International Affective Picture Series (IAPS)

During the training phase (Figure 1A), participants learned the 60 face-picture pairs as they were displayed, one pair at a time, on a computer monitor for 3.5 seconds followed by a 0.5-second fixation cross. The cue faces (225 x 225 pixels) were displayed on the left side of the screen, and the neutral or negative target images (225 x 225 pixels) on the right side of the screen (Figure 1A, Panel 1). The pictures were displayed centered vertically on the computer screen and horizontally on either side of the midpoint such that the left edge of the face picture coincided with the left edge of the screen and the right edge of the other picture coincided with the right edge of the screen. Participants viewed subsets of 20 pairs at a time in different random order, and the three subsets were cycled through 3 separate times, with a recognition test at end of each subset of 20 pairs. Each set of 20 pairs had an equal proportion of female/neutral, female/negative, male/neutral, and male/negative face-picture pairs.

During the recognition test (Figure 1A, Panel 2), participants were shown the cue faces alongside two pictures: one that was originally paired with it and a consistent distractor picked randomly from the stimuli set that remained constant for all 3 recognition cycles. Each picture in the 20 pair subset, therefore, appeared once as a target and once as a distractor in the recognition test. The cue faces (225 x 225 pixels) were centered vertically on the right side of the screen such that the right edge of the picture coincided with the right edge of the screen. Both the target and distractor pictures (225 x 225 pixels) were stacked vertically on the left side of the screen such that the left edge of the pictures coincided with the left edge of the screen. The participants were asked to identify the correct face-picture association by pressing the “T” or “B” button on the response box if the correct target image was on top or bottom of the screen. The intertrial interval presented a grey (neutral) fixation cross for 0.5 seconds. For each participant, each set of 20 pairs was cycled through three times to overtrain them on the pairs as previous research indicated that the average number of training cycles required to learn the face-picture associations is close to two (M= 1.76, SD=0.61; Depue, Banich, & Curran, 2006). Participants with lower than 90% on the third learning cycle were removed prior to data analysis. After the learning phase, participants were fitted with a 40-channel electrode cap.

During the experimental phase, the participants were comfortably seated in the testing room while their EEG was recorded. The participants viewed 40 of the 60 cue faces (225 x 225 pixels) centered vertically and horizontally on the computer screen. Half the faces were from the think condition and half the no-think condition. For both conditions, a trial consisted of a face framed by a colored border (30 x 30 mm) presented on a computer screen for 3.5 seconds followed by 0.5 second intertrial interval represented by a grey fixation cross. The border color was varied across trials to signal which strategy a participant should use: green for think trials and red for no-think trials (Figure 1B). For think trials, participants were instructed to concentrate on the memory of the target picture previously associated with the cue face, or for no-think trials, to try to prevent recall of the previously associated picture. For each condition (think/no-think), participants viewed 20 faces 10 times each. After every 2 cycles, a one-minute break was given to the participants to rest their eyes. Of the original 60 pairs, 20 were assigned to a baseline condition and not shown in the experimental phase.

During the final recall phase (Figure 1C), participants were shown each of the 60 faces (225 x 225 pixels) for 3.5 seconds each followed by an intertrial interval of 0.5 second represented by a grey fixation cross. Between each cue face stimulus, they were asked to describe the correct target image in two to three words. Their verbal responses were recorded by the researcher and provided a behavioral measure of cued recall accuracy to assess suppression effects. These descriptions were then scored correct or incorrect by two independent judges (inter-rater reliability was .98). Differences in scores allotted by these two independent judges were adjudicated by a third blind judge. Following the completion of the final test phase, the participant was debriefed.

### 2.4 EEG Recording and Analysis

Continuous EEG was recorded from 40 Ag/AgCl electrodes embedded in a 40-channel Neuroscan Quik-Cap. Electrical Oculogram (EOG) was recorded by additional electrodes positioned above and below the left eye (vertical movements), and on the outside edge of the right and left eye (horizontal movements). An additional reference electrode was positioned on the electrically neutral tip of the nose. EEG signals were amplified by a 40-channel Neuroscan NuAmps amplifier at a sample rate of 500 samples per second. Electrode impedances of 5 kΩ were obtained at all active sites. Data acquisition and post-acquisition processing was performed using Scan 4.3 software. Continuous EEG was filtered off-line with a bandwidth of 0.1 to 70 Hz, with a gain of 19. Continuous EEG recordings were partitioned into epochs (−200–850 ms) time-locked with the presentation of each face cue and baseline corrected using the −200 to 0 ms time window as baseline. Epochs were manually inspected and marked as bad in the presence of overwhelming electrical artifact, and epochs with observed potentials outside the ±70 μV range were automatically rejected. The remaining epochs were averaged by group based on the cross of valence (negative vs. neutral) by cognitive strategy (think vs. no-think).

Four target electrode sites of interest were chosen to be consistent with previous ERP studies of memory suppression (e.g., Bergstrom et al., 2007, 2009; Chen et al., 2012): electrodes F3 and F4 (left and right frontal sites, respectively) and electrodes P3 and P4 (left and right parietal sites, respectively). Averaged ERP waveforms for each of the 4 stimulus conditions, at each electrode of interest, were visually inspected and time windows for assessment of the parietal P1 (100-150 ms), frontal P3 (350-450 ms), and parietal LC (450-550 ms) were identified. Mean area under the curve (equivalent to mean amplitude as areas under the baseline are given negative values) were computed for each participant, for each experimental condition, at each electrode of interest and timewindow of interest. There was no visually discernable N2 (typically peaks at 200 ms post-stimulus) effect at the frontal sites of interest, this ERP component was not analyzed.

## 3 Results

### 3.1 Recognition Accuracy

Recognition accuracy scores derived from behavioral responses were calculated for the total training phase and for each of the three subdivisions. Overall mean recognition accuracy across all participants and repetitions was 96.2% (*SD* = 3.8%), indicating that participants succeeded in learning stimulus pairings. As expected, mean recognition accuracy was lowest after the first training block (*M* = 91.9%, *SD* = 9.3%), but was similar across the second (*M*= 98.5%, *SD* = 1.9%) and final (*M*= 98.4%, *SD*=1.8%) training blocks.

### 3.2 Rumination Scores

Mean Total RRS score for all participants was 37.65 (*SD* = 8.94) on a scale of 22 to 88. On a scale of 5 to 20, the mean score for all participants on the Reflection subscale was 9.48 (*SD* = 3.48) and on the Brooding subscale was 8.65 (*SD* = 2.21).

### 3.3 Behavioral Data

Recorded verbal responses on the cued recall test phase were scored correct or incorrect by two independent judges (inter-rater reliability was .98). Differences in scores allotted by these two independent judges were adjudicated by a third blind judge. Baseline-corrected recall scores provided behavioral data for analyses (Figure 2). Total recall scores were corrected for baseline learning by substracting the baseline recall rate for the 20 baseline stimuli, controlling for valence, from the observed recall rate for the experimental conditions (think and no-think). Baseline scores reflect the amount of forgetting that could be expected from passive memory decay over the course of the study. The scores were analyzed using a 2 valence (neutral vs. negative) x 2 strategy (think vs. no-think) ANOVA, with both factors manipulated within subjects. Results showed that there was a main effect of valence with more items being recalled above the baseline in the neutral condition (*M* = 21.9%, *SE* = 3.1%) than the negative condition (*M* = 6.3%, *SE* = 4.4%), *F* (1, 21) = 9.84, *p* < .01, partial η^2^ = .32. A main effect of strategy was also evident as there was a significant difference between the number of items recalled above the baseline in the think (*M*= 20.1%, *SE* = 3.4%) versus no-think conditions (*M*= 8.1%, *SE* = 2.9%), *F*(1, 21) = 22.66, *p* < .01, partial η^2^ = .52. However, there was no significant interaction effect observed (*p* >.19). The think vs no-think comparison was significant for each valence in isolation (p’s < .01). Moreover, the baseline corrected neutral no-think mean percent correct was significantly positive (p = .03), but the baseline corrected mean negative no-think mean percent correct was not significantly different from 0 (p > .50).

**Figure 2.**
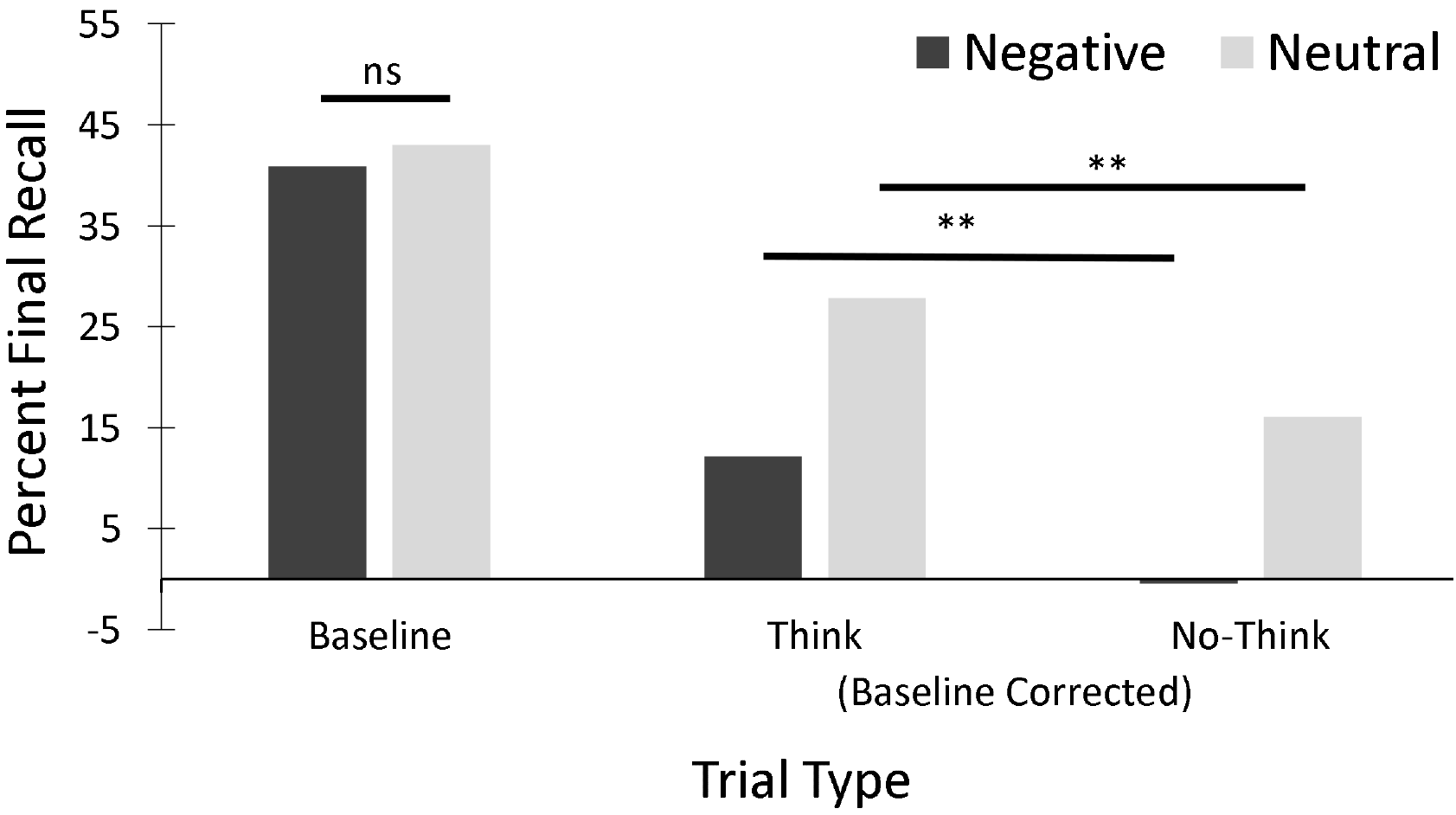
Mean final cued recall percent for negative and neutral baseline face-picture pairs (on left) and negative and neutral think and no-think pairs. Bars indicate valence was not significant for baseline pairs, but strategy (think vs. no-think) was significant (** p < .01) for both neutral and for negative pairs. Think and no-think means have been baseline corrected. The main effect of strategy and of valence are both significant (p < .002) for the baseline corrected results (on right), but the interaction was not significant.

Because neither of the no-think conditions yielded significantly negative baseline corrected means, there was no significant memory suppression effect for no-think trials. However, there was a significant think/no-think differential, indicating that participants were able to successfully halt cued retrieval on no-think trials more often than for the think trials.

### 3.4 ERP Data

For each individual, ERP results to face cue images were quantified by area under curve measures at each electrode site of interest (F3, F4, P3, & P4) for each combination of valence by strategy at time windows of interest (P1, 100-150 ms, P3, 350-450 ms, LC, 450-550 ms). The area measures were submitted to 2 valence (negative, neutral) x 2 strategy (think, no-think) x 2 laterality of electrode site (left, right) repeated measures ANOVAs. ERP waveforms at the four electrode sites of interest are depicted in Figure 3. Scalp plots of the voltage effects of interest are presented in Figure 4. The left panel presents the negative minus neutral trial voltage difference in the P1 (100-150 ms) time window, and indicates a primarily posterior scalp distribution. The middle panel presents the think minus no-think voltage difference in the P3 (350-450 ms) time window and indicates distinct frontal and posterior foci. The right panel presents the think minus no-think voltage difference in the LC (450-550 ms) time window and indicates a primarily posterior scalp distribution.

**Figure 3.**
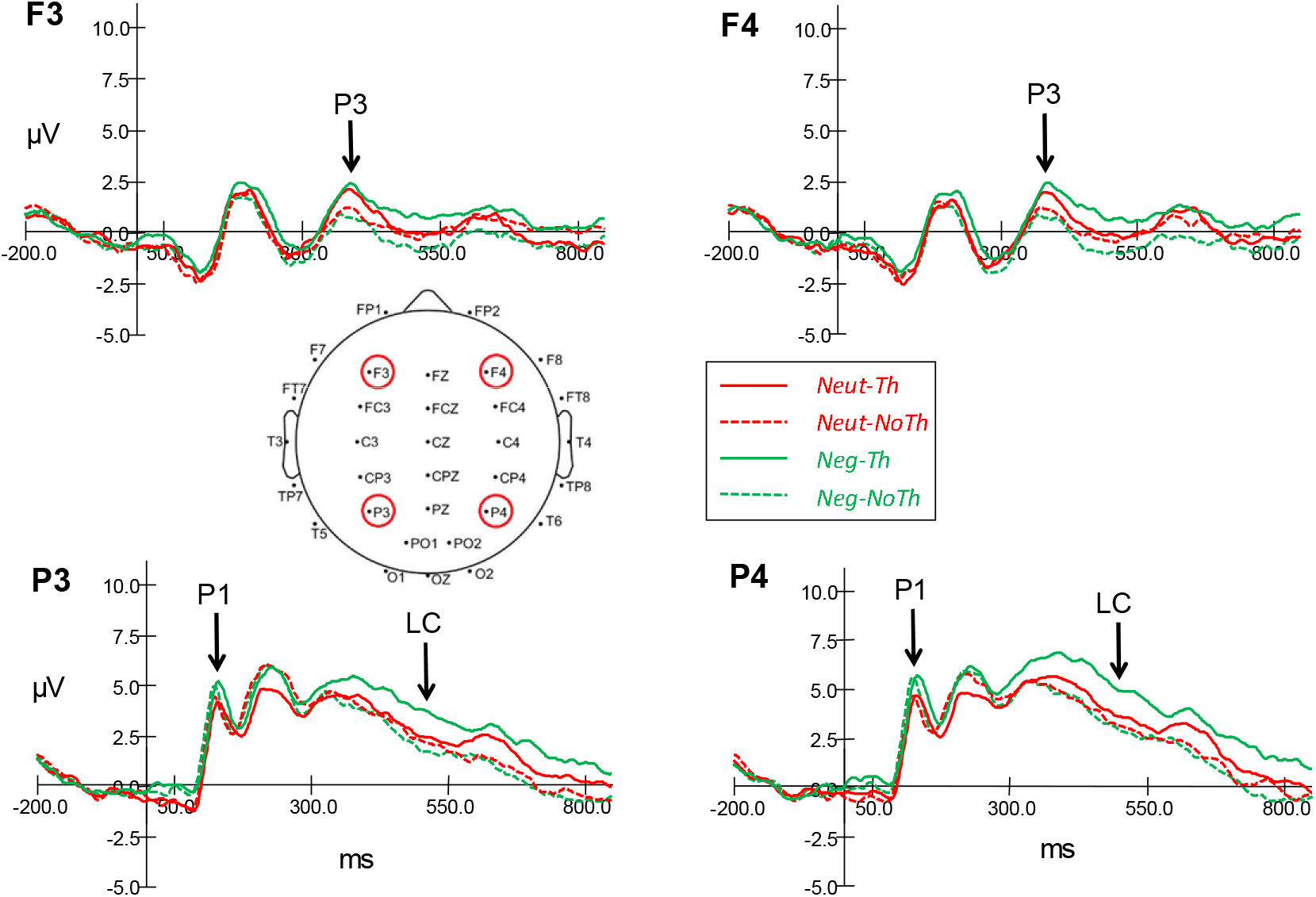
Grand mean ERPs for the four experimental conditions (neutral-think, neutral-nothink, negative-think, negative-nothink) at all four electrode sites (F3-left frontal, F4-right frontal, P3-left parietal, P4-right parietal).

**Figure 4.**
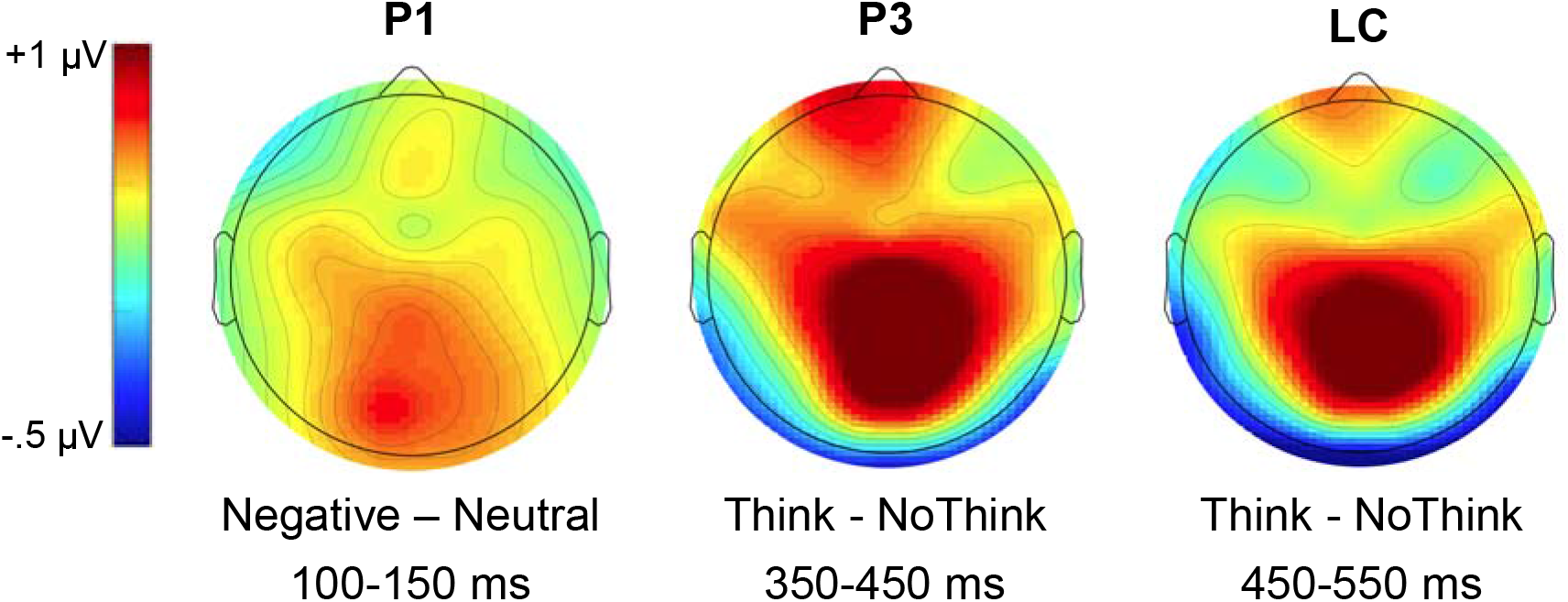
Scalp distribution for comparisons of interest in each time window of interest: P1 (100-150 ms), P3 (350-450 ms), LC (450-550 ms).

#### 3.4.1 Posterior P1 (100-150 ms)

The posterior P1 is thought to reflect attention to sensory characteristics of the stimuli (e.g., face cue). There was a main effect of valence with a greater positive mean ERP area in the negative condition (*M* = 198.2, *SE* = 35.7) than the neutral condition (*M* = 162.0, *SE* = 33.5), *F*(1, 22) = 9.17, *p* =.006, partial η^2^ = .29. There were no other significant interaction effects observed (all F’s < 3.04 and *p*’s > .09).

#### 3.4.2 Frontal P3 (350-450 ms)

The frontal P3 results demonstrated emergence of ERP activity associated with the exercise of conscious control. There was a main effect of strategy with a greater positive mean ERP area in the think condition (*M* = 158.0, *SE* = 57.2) than the no-think condition (*M* = 61.1, *SE* = 39.9), *F*(1, 22) = 7.32, *p* = .013, partial η^2^ = .25. No other main effects or interactions were significant.

#### 3.4.3 Posterior LC (450-550 ms)

The posterior LC is thought to be related to retrieval success and to further processing of retrieved target (Bergman et al., 2007, 2009; Chen et al., 2012; Rugg, 1995). There was a main effect of strategy with a greater positive mean ERP area in the think condition (*M* = 381.1, *SE*= 60.7) than the no-think condition (*M* = 261, *SE* = 56.4), *F(1*, 22) = 4.56, *p* = .044, partial η^2^ =.17. There was a main effect of laterality as there was a larger positive mean ERP area on the right electrode site (*M* = 375.7, *SE* = 51.0) compared to the left (*M* = 266.8, *SE* = 58.6), *F*(1, 22) = 8.19, *p* = .009, partial η^2^ = .27. There was also an interaction pattern between valence and strategy (see Figure 5) was also continued, *F*(1, 22) = 4.64 *p* = .043, partial η^2^ = .17. Further analysis of the strategy effect at each level of valence revealed a significantly larger area under the ERP for think than for no-think trials for the negatively valenced pictures, *F*(1, 22) = 9.89, *p* = .005, partial η^2^ = .31, but not for the neutral valenced pictures (p > .05).

**Figure 5.**
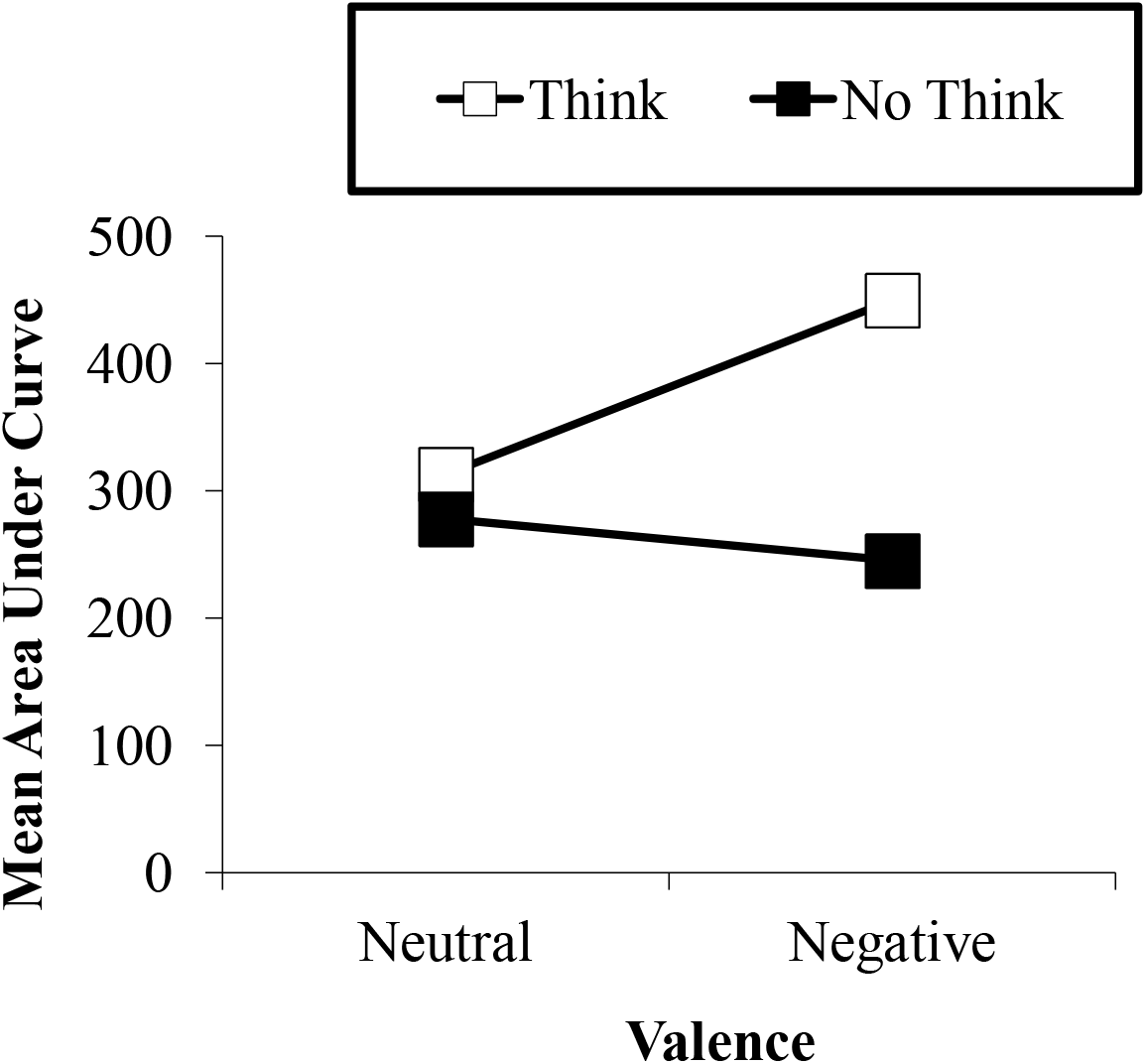
Mean area under ERP for posterior LC (450-550 ms post-cue), broken down by valence and cognitive strategy.

### 3.5 Correlational Analyses with Rumination Scores

In order to explore if rumination scores were significantly related to memory inhibition, the data was analyzed using Pearson correlations. We compared rumination (all three rumination scores: Total RRS, negative Brooding, & positive Reflection) with behavioral recall in the nothink category, strategy differences in behavioral recall between think and no-think categories, and area measures for Window 5 (LC) at parietal electrodes (i.e., P3 & P4) for no-think trials and strategy (think-minus-no-think) differences. We also added post-hoc correlations between rumination scores and final cued recall for think and no-think trials in isolation (see Table 1 for correlations) to allow for better interpretation of the planned correlations. There was a positive correlation between RRS-Brooding and the combined (across electrodes P3 & P4) posterior LC ERP think/no-think difference, *r*(23) = .48, *p* = .021. This correlation is opposite in direction to our prediction (see Discussion for more). There was also a nonsignificant positive correlation, *r*(23) = .30, *p* > .05 between RRS-Brooding and the final cued recall think/no-think difference (see Discussion).

**Table 1.**
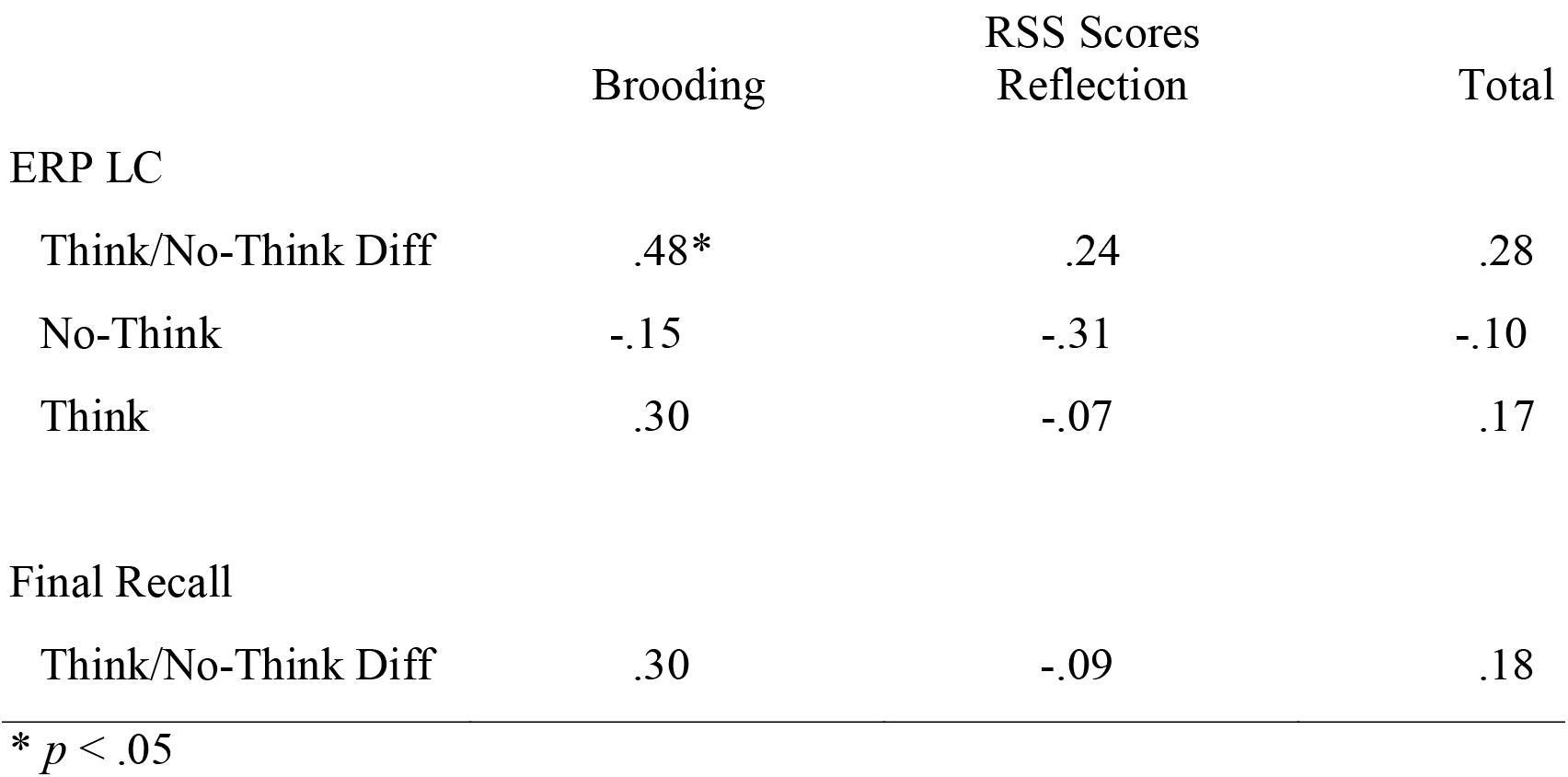
Pearson’s correlation between rumination and the posterior LC, and baseline final recall (N=23)

## 4 Discussion

Previous research has indicated that control mechanisms in the brain can be activated to actively suppress completion of a memory retrieval initiated by a memory cue (Anderson & Green, 2001; Bergstrom et al., 2007; Depue et al., 2007), and repeated interruption of cued retrieval may eventually weaken the ability of a cue to drive retrieval of an associated target (Anderson & Levy, 2009; Levy & Anderson, 2002) resulting in a long-term suppression of memory, but this effect is not always observed (e.g., Bulevich et al., 2006). It also appears that cognitive control of memories differs depending on whether they contain information with negative or neutral emotional content. People, seemingly, do a better job of suppressing stimuli that is negative in content as compared to more neutral stimuli (e.g., Depue et al., 2006; Depue et al., 2013; Lambert et al., 2010).

The purpose of the present study was to use ERP methodology to replicate and augment prior research findings (Bergstrom et al., 2007, 2009; Chen et al., 2012; Hanslmayr et al., 2009; Zhang et al., 2016) on memory suppression during the think/no-think task. An additional goal was to see if the relationship between rumination and memory suppression indexed by cued recall measures (Dieler et al., 2014; Fawcett et al., 2015) would also be found between rumination and neurophysiological ERP markers associated with memory suppression.

Previous ERP studies of face perception have found an early positivity (P1) over posterior scalp locations peaking 100-150 ms post-presentation (Olivares, Iglesias, Saavedra, Trujillo-Barreto, Valdes-Sosa, 2015). We observed a posterior P1 (100-150 ms) that was greater for negative than for neutral trials during the think/no-think phase. By contrast, the posterior P1 has been found to be more positive for think than for no-think trials in one previous study (Chen et al., 2012) that used emotionally valenced target scenes and neutral face cues. Our finding that the posterior P1 during neutral face presentation is modulated by emotional valence of an associated item is, to our knowledge, novel in the literature. However, there is a growing appreciation in the literature on face perception that the P1 is sensitive to emotional expressions of faces (e.g., Earls, Curran, & Mittal, 2016). Take together our results, combined with those of Chen et al. (2012), suggest that early visual attention may be modulated both by retrieval strategy and by quick partial retrieval of emotional content associated with the current visual stimulus.

Both Chen et al. (2012) and Zhang et al. (2016) reported more negative N2 (~200 ms peak) for no-think than for think trials at frontal sites. The present study failed to observe a discernable frontal N2 effect (see Figure 3). Previous research on the think/no-think task has indicated participants may vary in their cognitive control strategies (van Schie et al., 2013) with active memory suppression and mental substitution (try to think about something else during nothink trials) being the 2 major ones. The frontal N2 has been argued to be related to a suppression strategy (Bergstrom et al., 2009), indicating that our participants may have used substitution rather than active suppression more often than the participants in these other studies. Our instructions encouraged maintenance of eye fixation on the face cue and to either allow (think) or not allow (no-think) the associated picture to come to mind, but did not provide more specific instructions on how to do this. Another factor that may have reduced our ability to observe a frontal N2 was our use of a nasal reference electrode location as opposed to the more common combined left/right mastoids (behind the ear).

Our results yielded a frontal P3 (positivity, 350-450 ms) that was greater for think than for no-think trials. Hanslymayr et al. (2009) reported evidence that this P3-like effect acted as an alerting signal for cognitive control of memory during the think/no-think task using neutrally valenced face-word pairs. Our use of emotionally valenced picture targets allowed us to look for the influence of emotion processing on this ERP component. However, an interaction of emotional valence and memory strategy was not observed in our study until a later (450-550 ms time window). Our results suggest that the frontal P3 alerting signal is not directly influence by emotional processing during the think/no-think task.

The present study found an interaction of emotional valence and memory strategy for the posterior LC (450-550 ms), such that think trials had a greater positive area than no-think trials, and this think/no-think difference was greater for negative than neutral trials. In fact, only the negative trials yielded a significant think-minus-no-think difference. Zhang et al. (2016) found a similar interaction using a similar set of neutral face cues and emotionally valenced target pictures. However, the think-minus-no-think difference was significant for both negative and neutral trials. Of interest to our results is the group comparison reported in this study. Zhang and colleagues also reported a significant group difference in this interaction such that the strategy effect was much greater for negative trials than for neutral trials in a group of depressed individuals. Whereas the non-depressed group yield similar strategy effects for both negative and neutral trials. Consideration of our finding that RSS-Brooding scores were positively correlated with the think vs no-think strategy effect in the posterior LC (450-550 ms) in conjuction with the results of Zhang et al. (2016) suggests that rumination acts to increase the posterior LC for negative trials. The pattern of the data in both studies suggests a reduction in cognitive control over negative think trials in particular, perhaps due to perseverative focus on negative think targets.

Depue et al. (2007) conducted an fMRI study of face-picture pairs and reported evidence for a 2-control process strategy, with an emotional control network activated for valenced material in addition to the control network activated for think and no think condition. More recently, Gagnepain et al. (2017) found evidence for dorso-lateral prefontal control over the hippocampal complex and amygdala via distinct pathways, they also found evidence of profrontal control over posterior visual cortex as well as from temporal lobe. Our results documented earlier strategy modulated effects at frontal sites (frontal P3), as well as later posterior interaction of valence and strategy (posterior LC).

Another goal of this study was to observe the role of ruminative styles of thinking on memory suppression as measured by the RRS (Nolen-Hoeksema & Morrow, 1991). This provides a clinically useful corrolarly between this study and management of negative memories. We predicted that high ruminators would not be able to exert memory control processes as efficiently as low ruminators resulting in smaller differences in scores across the four experimental conditions for the former. Results, however, indicated the opposite relationship between RRS scores and ERP suppression effects. The only significant associations found in the analyses were positive correlations between Brooding subscale scores and the combined (across electrodes P3 & P4) posterior LC ERP think/no-think difference thought to index retrieval success (e.g. Curran et al., 2006). This relationship was contrary to what we expected and suggests that high brooders show better suppression of presented material. This relationship also contradicts Hertel and Gerstle’s (2003) findings demonstrating a negative correlation between suppression effect and depressive symptoms, specifically depressive rumination. One explanation for this pattern is that, since our sample consisted of college students who were not necessarily clinically depressed, the high scorers on the brooding subscale probably do not capture maladaptive patterns of brooding in our study. Instead, it is likely that we are observing a pattern of individuals engaging in moderate levels of rumination being most adept at actively suppressing memories. Similar findings of a relationship between higher reported trauma was found with better suppression scores in non-PTSD patients (Hulbert and Anderson, 2018), while individuals diagnosed with clinical levels of PTSD demonstrate poorer memory suppression (Catarino et al., 2015). This might suggest that moderate (and not severe) levels of aversive experiences may make individuals better at suppressing negative stimuli due to prior experiences of doing so or a process of resilience. Further research would, therefore, be warranted to examine if specifically training patient groups on memory control strategies in conjunction with distress reduction of trauma or mood symptoms would improve their suppression of unwanted past memories.

## 5 Conclusion

In summary, the present study provides further evidence for think/no-think related ERP effects previously reported in the literature: (a) a frontal P3 (350-450 ms) alerting signal in preparation of cognitive control of memory retrieval modulated by strategy but not emotional valence, (b) a later posterior LC (450-550 ms) modulated by a combination of strategy and emotional valence. The present study extends evidence regarding an early posterior P1 by finding that it is modulated by emotional valence, not memory strategy as in earlier reports (e.g., Chen et al., 2012). Our results also provide the first report of a relationship between rumination and the posterior LC thought to index retrieval success (e.g. Curran et al., 2006). Considering that this study was able to establish these findings using a college population, the next step would be to research clinical populations using this paradigm. One direction would be to examine the role of depressive rumination in greater detail by including more diagnostic scales of depression in the design as well as testing a larger sample. Our current findings that suppression effects at parietal electrode sites are positively related to rumination indicate that suppression may possibly be a learned response. Another interesting future direction is to examine if memory suppression mechanisms are somehow disrupted for clinical disorders like PTSD and OCD that are characterized by intrusive thoughts, especially when the stimuli used is negatively valenced. As mentioned before, such research could have promising clinical implications as identifying such a deficit may propel research in the direction of finding ways to teach individuals to enhance cognitive control mechanisms before their symptoms become debilitating.

## Data Availability

Data available upon request

## Acknowledgments

Our sincerest thanks to Brendan Depue for sharing his stimuli set with us and providing guidance in setting up the experimental paradigm. Our special thanks to all the participants who put forth their time towards our research efforts.

## Conflict of Interest

The authors do not have any potential conflict of interests to disclose.

## Author contributions

*Conceptualization* – AN, MF; *Methodology* – AN, JE, MF; *Investigation* – AN, JE; *Formal Analysis* – AN, JE; *Resources* – MF; *Writing* – AN, MF; *Visualization* – AN, JE; *Supervision* – MF.

## Notes

### Competing Interest Statement

The authors have declared no competing interest.

### Funding Statement

No external funding was received for this study.

## REFERENCES

Anderson, M.C., & Green, C. (2001). Suppressing unwanted memories by executive control. Nature, 410, 366–369. https://doi.org/10.1038/35066572

Anderson, M.C., & Levy, B.J. (2007). Theoretical issues in inhibition: Insights from research on human memory. In: D.S. Gorfein & C.M. MacLeod (Eds.), Inhibition in Cognition (pp. 81-100). Washington, DC: American Psychological Association. https://doi.org/10.1037/11587-005

Anderson, M., & Levy, B. (2009). Suppressing Unwanted Memories. Current Directions in Psychological Science, 18, 189–194. https://doi.org/10.1111/j.1467-8721.2009.01634.x

Bergstrom, Z.M., Velmans, M., Fockert, J., & Richardson-Klavehn, A. (2007). ERP evidence for successful voluntary avoidance of conscious recollection. Brain Research, 1151, 119–133. https://doi.org/10.1016Zj.brainres.2007.03.014

Bergstrom, Z.M., Fockert, J., & Richardson-Klavehn, A. (2009). ERP and behavioural evidence for direct suppression of unwanted memories. NeuroImage, 48, 726–737. https://doi.org/10.1016/j.neuroimage.2009.06.051

Bulevich, J., Roediger, H., Balota, D., & Butler, A. (2006). Failures to find suppression of episodic memories in the think/no-think paradigm. Memory & Cognition, 34, 1569–1577. https://doi.org/10.3758/BF03195920

Catarino, A., Kupper, C.S., Werner-Seidler, A., Dalgleish, T., & Anderson, M.C. (2015). Failing to forget: Inhibitory-control deficits compromise memory suppression in posttraumatic stress disorder. Psychological Science, 26, 604–616. https://doi.org/10.1177/0956797615569889

Chen, C., Liu, C., Huang, R., Cheng, D., Wu, H., Xu, P., Mai, X., & Luo, Y. (2012). Suppression of aversive memories associates with changes in early and late stages of neurocognitive processing. Neuropsychologia, 50, 2839–2848. https://doi.org/10.1016/j.neuropsychologia.2012.08.004

Curran, T., Tepe, K.L., & Piatt, C. (2006). ERP explorations of dual processes in recognition memory. In: Zimmer, H.D., Mecklinger, A., & Lindenberger, U. (Eds.), Binding in Human Memory: A Neurocognitive Approach. Oxford University Press, Oxford. https://doi.org/10.1093/acprof:oso/9780198529675.001.0001

Dieler, A., Herrmann, M., & Fallgatter, A. (2014). Voluntary suppression of thoughts is influenced by anxious and ruminative tendencies in healthy volunteers. Memory, 22(3), 184–193. https://doi.org/10.1080/09658211.2013.774420

Depue, B.E., Banich, M.T., & Curran, T. (2006). Suppression of emotional and non-emotional content in memory: Effects of repetition on cognitive control. Psychological Science, 17, 441–447. https://doi.org/10.1111/j.1467-9280.2006.01725.x

Depue, B.E., Curran, T., & Banich, M.T. (2007). Prefrontal regions orchestrate suppression of emotional memories via a two-phase process. Science, 317, 215–219. https://doi.org/10.1126/science.1139560

Depue, B.E., Ketz, N., Mollison, M.V., Nyhus, E., Banich, M.T., & Curran, T. (2013). ERPs and Neural Oscillations during volitional suppression of memory retrieval. Journal of Cognitive Neuroscience, 25, 1624–1633. https://doi.org/10.1162/jocn_a_00418

Duzel, E., Vargha-Khadem, F., Heinze, H., & Mishkin, M. (2001). Brain activity evidence for recognition without recollection after early hippocampal damage. Proceedings of the National Academy of Sciences of the United States of America, 98, 8101–8106. https://doi.org/10.1073/pnas.131205798

Earls, H., Curran, T., & Mittal, V. (2016). Deficits in Early Stages of Face Processing in Schizophrenia: A Systematic Review of the P100 Component. Schizophrenia Bulletin, 42, 519–527. https://doi.org/10.1093/schbul/sbv096

Fawcett, J., Benoit, R., Gagnepain, P., Salman, A., Bartholdy, S., Bradley, C., Chan, D. K.; Roche, A.; Brewin, C. R.; Anderson, M. (2015). The origins of repetitive thought in rumination: Separating cognitive style from deficits in inhibitory control over memory. Journal of Behavior Therapy and Experimental Psychiatry, 47, 1–8. https://doi.org/10.1016/j.jbtep.2014.10.009

Gagnepain, P., Hulbert, J., & Anderson, M.C. (2017). Parallel regulation of memory and emotion supports the suppression of intrusive memories. The Journal of Neuroscience, 27, 64236441. https://doi.org/10.1523/JNEUROSCI.2732-16.2017

Hanslmayr, S., Leipold, P., Pastotter, B., & Bauml, K.H. (2009). Anticipatory signatures of voluntary memory suppression. The Journal of Neuroscience, 29, 2742–2747. https://doi.org/10.1523/JNEUROSCI.4703-08.2009

Hertel, P.T., & Gerstle, M. (2003). Depressive deficits in forgetting. Psychological Science, 14, 573–578. https://doi.org/10.1046/j.0956-7976.2003.psci_1467.x

Hulbert, J. C., & Anderson, M.C. (2018). What doesn’t kill you makes you stronger: Psychological trauma and its relationship to enhanced memory control. Journal of Experimental Psychology: General, 147, 1931–1949. http://dx.doi.org/10.1037/xge0000461

Koster, E., De Lissnyder, E., Derakshan, N., & De Raedt, R. (2011). Understanding depressive rumination from a cognitive science perspective: The impaired disengagement hypothesis. Clinical Psychology Review, 31, 138–145. https://doi.org/10.1016/jxpr.2010.08.005

Lang, P.J., Bradley, M.M., & Cuthbert, B.N. (1995). The International Affective Picture System (IAPS). Gainesville: University of Florida, Center for Research in Psychophysiology.

Lambert, A.J., Good, K.S., & Kirk, I.J. (2010). Testing the repression hypothesis: Effects of emotional valence on memory suppression in the think-No think task. Consciousness and Cognition, 19, 281–293. https://doi.org/10.10167j.concog.2009.09.004

Levy, B.J., & Anderson, M.C. (2002). Inhibitory processes and the control of memory retrieval. Trends in Cognitive Sciences, 6, 299–305. https://doi.org/10.1016/S1364-6613(02)01923-X

Mclaughlin, K., & Nolen-Hoeksema, S. (2011). Rumination as a transdiagnostic factor in depression and anxiety. Behaviour Research and Therapy, 49(3), 186–193. https://doi.org/10.1016Zj.brat.2010.12.006

Nolen-Hoeksema, S., & Morrow, J. (1991). A prospective study of depression and posttraumatic stress symptoms after a natural disaster: The 1989 Loma Prieta earthquake. Journal of Personality and Social Psychology, 61, 115–121. https://doi.org/10.1037/0022-3514.61.1.115

Nolen-Hoeksema, S. (2000). The role of rumination in depressive disorders and mixed anxiety/depressive symptoms. Journal of Abnormal Psychology, 109, 504–511. https://doi.org/10.1037/0021-843X.109.3.504

Oldfield, R. C. (1971). The assessment and analysis of handedness: The Edinburgh Inventory. Neuropsychologia, 9, 97–113. https://doi.org/10.1016/0028-3932(71)90067-4

Olivares, E.I., Iglesias, J., Saavedra, C., Trujillo-Barreto, N.J., Valdes-Sosa, M. (2015). Brain signals of face processing as revealed by event-related potentials. Behavioural Neurology, 2015: 514361. https://doi.org/10.1155/2015/514361

Rugg, M.D. (1995). ERP studies of memory. In: Rugg, M.D., & Coles, M.G.H. (Eds.), Electrophysiology of Mind (pp. 133–170). Oxford University Press, Oxford.

Sacchet, M.D., Levy, B.J., Hamilton, J.P., Maksimovskiy, A., Hertel, P. T., Joormann, J., Anderson, M.C., Wagner, A.D., & Gotlib, I.H. (2017). Cognitive and neural consequences of memory suppression in major depressive disorder. Cognitive, Affective, & Behavioral Neuroscience, 17, 77–93. https://doi.org/10.3758/s13415-016-0464-x

Treynor, W., Gonzalez, R., & Nolen-Hoeksema, S. (2003). Rumination reconsidered: A psychometric analysis. Cognitive Therapy & Research, 27, 247–259. https://doi.org/10.1023/A:1023910315561

Van Schie, K., Geraerts, E., & Anderson, M.C. (2013). Emotional and non-emotional memories are suppressible under direct suppression instructions. Cognition & Emotion, 27, 1122–1131. https://doi.org/10.1080/02699931.2013.765387

Zhang, D., Xie, H., Liu, Y., & Luo, Y. (2016). Neural correlates underlying impaired memory facilitation and suppression of negative material in depression. Sci Rep, 6, 37556. https://doi.org/10.1038/srep37556

